# An exploration of attributes effecting the relationship between competencies and performance of CHWs in a rural block

**DOI:** 10.1101/2022.01.11.22268912

**Authors:** Revadi G, Ankur Joshi, Abhijit P Pakhare

## Abstract

**Introduction:** Induction of Community Health Workers (CHWs) into various health programs is considered as an integral strategy to achieve universal health coverage. Thus, it is prudent to explore and understand the CHWs individual and the system level interactions and their further translation into performance and actionable domains.

**Objective:** To quantify the relationship structure between envisioned competencies and CHW performance with factors operating at individual and the immediate system level as effect modifiers.

**Methodology:** A cross-sectional study was done in primary rural health care settings of Madhya Pradesh state in Central India. CHWs were stratified as relatively low performing (RLP) and relatively high performing (RHP) based on their annual performance-based incentives for the year (April 2017- March 2018). CHWs were administered a self-reported questionnaire that included socio demographic details, knowledge, skills assessment and availability of logistics.

**Results:** Among the 90 eligible CHWs, 31 RLP and 30 RHP CHWs were interviewed. The CHWs performance was found to be significantly associated with age, education, caste, presence of under 5 children, knowledge and competency scores. However, while adjusting for the confounders only age and educational status were found to be significant. Further cluster analysis revealed two clusters based on individual and system characteristics which was found to influence the CHWs performance.

**Conclusion:** The relationship between the CHWs truest competencies and performance based incentivisation tends to have been influenced by age and education which could help in developing a more focused supportive supervision catering to their needs.

## Introduction

World Health Organization defines “Community health workers as those who are representatives selected by and answerable to the community, they work for with support from the health system with shorter training than professional health workers” [1,2]. One of the key components of the National Health Mission was to provide every village in India with a trained female community health activist-CHW [3] .CHWs were primarily involved in service delivery, referral of sick patients, conducting surveys, community mobilization and advocacy activities[4]. NHM in India had developed a performance-based incentives system (PBI) [5] wherein the CHWs were paid according to the health activities they perform under various National Programs (Supplementary file 1).

Proximate measures of CHWs performance includes knowledge, self-efficacy, job satisfaction and competency that occurs at the individual level of CHW. ^[6]^ Competency can be defined as degree to which the community health worker has the skills necessary to carry out the tasks assigned to him/her. ^[6]^ It was believed that competency was difficult to measure as it was dependant on theoretical understanding i.e, knowledge, or practical understanding i.e, training, work experience, support of supervisors, peers and community involvement.

This study was conducted with the primary objective to establish if there is a directional relationship between envisioned competencies and real-world performance (PBI)? Whether incentivization reflects in its truest sense the competency? which was translated to performance by the CHW. Moreover, what are those CHW characteristics at community level like population, distance to the health facility; health system level like availability of logistics, training and individual levels like age, education, caste, religion, marital status etc. ^[6,7]^ which may serve as effect modifiers in the aforesaid relationship. However, the existing literature showed almost negligible studies on CHW performance that could possibly answer this three-dimensional relationship. Hence this study was conducted in the rural setting of high focus state in developing country like India where CHWs are deployed in different health programs to achieve Universal health coverage.

## Methodology

### Study setting and Design

This cross-sectional study was conducted in a rural block (Obedullaganj, District Raisen) of Madhya Pradesh in Central India considering the operational feasibility. There were overall 213 CHWs (as of 2018) that included 189 in rural areas and 24 in the urban areas of the block.

### Study participants and study period

CHWs of Obedullaganj block were selected based on their performance in the year 2017-18. Data on PBI was retrieved from the Block Programme Management unit for the year (April 2017 – March 2018). The population of each of the village/catchment area served by the 189 CHWs in Obedullaganj block was retrieved from the ASHA (CHW database) of which 174 data were available excluding those in the urban areas. Villages were classified into three groups based on these population tertiles. Annual PBI of the financial year 2017-18 formed the basis of sample selection. From each group based on the population, all those CHWs who received above 75^th^ percentile of annual PBI were considered as Relatively High Performing (RHP) and those who received below 25^th^ percentile was considered as Relatively Low Performing (RLP).

The descriptive statistics of annual performance-based incentive received by CHWs has been summarised and its stratification by population group are displayed in Supplementary table 2. Out of 174 CHWs in Obedullaganj block, 90 were eligible for participation in the study after adjusting for the population, where only 61 participated in the detailed knowledge, skills and other assessments as shown in Supplementary Figure 1.

## Study variables and tools

### Socio-demographic details

Initially a questionnaire was developed for the CHWs (Supplementary file 2) containing name of the village, education, marital status, socio economic class, caste, religion, years of experience, trainings attended in the last 6 months, number of family members and number of under 5 children.

### Knowledge assessment tool

Secondly assessment of the knowledge in the form of 25 multiple choice questions adopted from the CHW training modules including various domains like maternal and child health, disease control, adolescent health was taken. Each correct answer was given a score of 1 and percentage of correct answers was calculated as knowledge score.

### Skills assessment tool

Skill assessment tool included checklist that contained home visit form with 16 steps, Home based neonatal care with 10 steps and Temperature measurement consisting of 9 steps adopted from the CHW training modules. There were asked to perform the steps as per their knowledge and it was filled against the checkbox by the investigator. For performance of every right step 1 mark was awarded leading to a maximum score of 35.

### Logistics supplies and drugs availability assessment tool

There was checklist of drugs, equipments and registers of CHWs that were cross verified. Excluding skill assessment and logistics checklist the remaining questionnaire was translated to the vernacular language.

### Data collection procedure

The questionnaire was deployed on paperless data collection platform Ona Inc. ^[8]^ The CHWs were approached for data collection at the nearest sub-Centre and in PHC and CHC during their monthly meetings with prior notice through their supervisor at least a week before the data collection. During the day of data collection, the CHWs were explained the purpose of the study with the help of Participant information sheet following which written informed consent was obtained.

## Ethics approval

This study protocol was reviewed and approved by Institutional Human Ethics Committee of AIIMS Bhopal (IHEC-LOP/2018/MD0027). Permissions were also obtained from Chief Medical Health Officer, Raisen and Block Medical Officer, Obedullaganj for data retrieval as well as for stakeholder interviews.

## Operational Definitions

1. ***National Health Mission:*** The NHM was launched by government of India in 2013 subsuming the National rural health and National Urban health mission. It envisages achievement of universal access to equitable, affordable and quality health care services that are accountable and responsible to people’s need. ^[9]^
2. ***Home based newborn care:*** CHWs are mandated to visit every newborn in her area with at least seven visits (Day 1, 3, 7, 14, 21, 28, 42) in case of home-based deliveries and six visits in case of Institutional deliveries (Day 3, 7, 14, 21, 28, 42). ^[10]^
3. ***Urban area:*** ^[11]^
  a. all places with a Municipality, Corporation or Cantonment or Notified Town Area
  b. all other places which satisfied the following criteria:
    I. minimum population of 5000
    II. At least 75% of the male working population was non-agriculture
    III. A density of population of at least 400 sq. Km. (i.e, 1000 per sq. Mile).
4. ***Rural area:*** Those areas that don’t follow the above definition will be included under rural India which accounts to 68.7%. ^[11]^
5. ***Village:*** In the rural areas the smallest area of habitation, viz., the village has a definite surveyed boundary, and each village is a separate administrative unit with separate village accounts. It may have one or more hamlets. The entire revenue village is one unit. ^[11]^
6. ***Block:* :** India is a large country comprising of 28 states and 8 union territories as per 2011 Census. These states and the union territories are divided into districts. Each district has 6 sub-divisions one of which is a block comprising of 80,000 to 1,00,000 population or 100 villages in total. ^[11]^
7. ***Village Health Sanitation and Nutrition Committee:*** Was introduced by National Rural Health Mission in 2005, to ensure community participation at all levels, which include participation as beneficiaries, in supporting health activities, in implementing, and even in monitoring and action-based planning for health programmes. ^[12]^
8. ***Sub centre*:** There should be 1 sub centre for every 5000 (plain area) & 3000 in (Hilly and tribal area) as per the norms of Indian Public Health Standards.
9. ***Primary Health Centre*** : There should be 1 PHC for every 30,000 (plain areas) & 20,000 in (Hilly and tribal areas).
10. ***Community Health Centre*:** Is the secondary level of contact between the community and the health facility. There should be 1 CHC for every 1,20,000 (plain area) & 80,000 in (Hilly and tribal area) as per the norms of Indian Public Health Standards.

## Data Analysis

Data in the form of excel sheet was imported from the Ona platform and following data cleaning data analysis was done using IBM SPSS version 24, R software version 4.1.0 with ggplot2 packages. Nominal or categorical variables were summarized as frequency and percentage. Continuous variables were summarized as mean and standard deviation when normally distributed and as median and interquartile range when non-normally distributed. For each potential determinant, association of nominal variable with the binary dependent outcome was done by Chi Square test, and of numerical variable by t-test or Mann-Whitney test as appropriately. Multivariate logistic regression was done to identify the independent determinants using adjusted odds ratios. Cluster analysis using two step method, was employed to identify the number and nature of clusters among the selected CHWs based on characteristics like age, education, performance group etc, P value < 0.05 was considered as significant.

## Results

The socio demographic details of the CHWs stratified according to their performance has been described in Table 1. Among which the median (IQR) age of CHW being 30(27-35) and the median(IQR) years of experience being 6(4-11). Variables like age (*P 0*.*001*), education (*P 0*.*006*), caste (*P 0*.*040*) and presence under 5 children (*P 0*.*013*) were found to be significantly associated by Chi square test.

**Table 1:** Association of CHWs performance with various levels of determinants (N =61)

| Sr<br>N0 | Variables | RLP<br>N=31 | RHP<br>N=30 | Total<br>N=61 | P value |
| --- | --- | --- | --- | --- | --- |
| **A** | **Individual Determinants** | **n (%)** | **n (%)** | **Total** | **P value** |
|  | **Socio-Demographic details** |  |  |  |  |
| 1 | Age |  |  |  |  |
|  | Less than or equal to 25 | 9(29) | 0 | 9 | 0.001 |
|  | More than 25 | 22(71) | 30(100) | 52 |  |
| 2 | Education |  |  |  |  |
|  | Up to Primary education | 18(58.1) | 7(23.3) | 25 | 0.006 |
|  | Others | 13(41.9) | 23(76.7) | 36 |  |
| 3 | Marital status |  |  |  |  |
|  | Married | 29(93.5) | 27(90) | 56 | 0.614 |
|  | Others | 2(6.5) | 3(10) | 5 |  |
| 4 | Religion |  |  |  |  |
|  | Hindu | 31(100) | 28(93.3) | 59 | 0.144 |
|  | Other religion | 0 | 2(6.7) | 2 |  |
| 5 | Caste |  |  |  |  |
|  | General | 3(9.7) | 7(23.3) | 10 | 0.040 |
|  | OBC | 12(38.7) | 11(36.6) | 23 |  |
|  | Scheduled Caste | 4(12.9) | 8(26.6) | 12 |  |
|  | Scheduled Tribe | 12(38.7) | 4(13.3) | 16 |  |
| 6 | Socio-economic status |  |  |  |  |
|  | Above poverty line | 15(48.4) | 10(33.3) | 25 | 0.232 |
|  | Below poverty line | 16(51.6) | 20(66.7) | 36 |  |
| 7 | No of family members |  |  |  |  |
|  | Less than or equal to 4 family members | 13(41.9) | 10(33.3) | 23 | 0.488 |
|  | More than 4 family members | 18(58.1) | 20(66.7) | 38 |  |
| 8 | Under 5 children |  |  |  |  |
|  | Less than or equal to two children | 13(41.9) | 10(33.3) | 23 | 0.013 |
|  | More than 2 children | 18(58.1) | 20(66.7) | 38 |  |
|  | **B Immediate systemic factors:** |  |  |  |  |
| 1 | Years of experience |  |  |  |  |
|  | <= 5 years of experience | 18(58.2) | 10(33.3) | 28 | 0.150 |
|  | > 5 years of experience | 13(41.9) | 20(66.7) | 33 |  |
| 2 | Training |  |  |  |  |
|  | Less than or equal to 5 | 30(96.8) | 26(86.7) | 56 | 0.053 |
|  | More than 5 | 1(3.2) | 4(13.3) | 5 |  |
|  | **Logistics availability** | **RLP**<br>**Median(IQR)** | **RHP**<br>**Median(IQR)** | **Total**<br>**Median(IQR)** | **P value** |
| 3.1 | Drug availability | 76.92<br>(57.7-84.6) | 69.23<br>(57.6-84.7) | 69.23<br>(57.6-84.6) | 0.648 |
| 3.2 | Equipment availability | 50<br>(30-70) | 60<br>(40-80) | 50<br>(30-80) | 0.408 |
| 3.3 | Registers availability | 100<br>(83.3-100) | 100<br>(83.3-100) | 100<br>(83-100) | 0.238 |

Table 2 shows that the overall knowledge (*P <0*.*001*), skills (*P 0*.*029*) and competency scores (*P <0*.*001*) were significantly associated with performance of CHWs following application of Unpaired t-test. Further, domain wise stratification of knowledge, skills and competencies showed that the difference in performance of RHP CHWs and the RLP CHWs was significant for all the domains of knowledge and competencies.

**Table 2:**
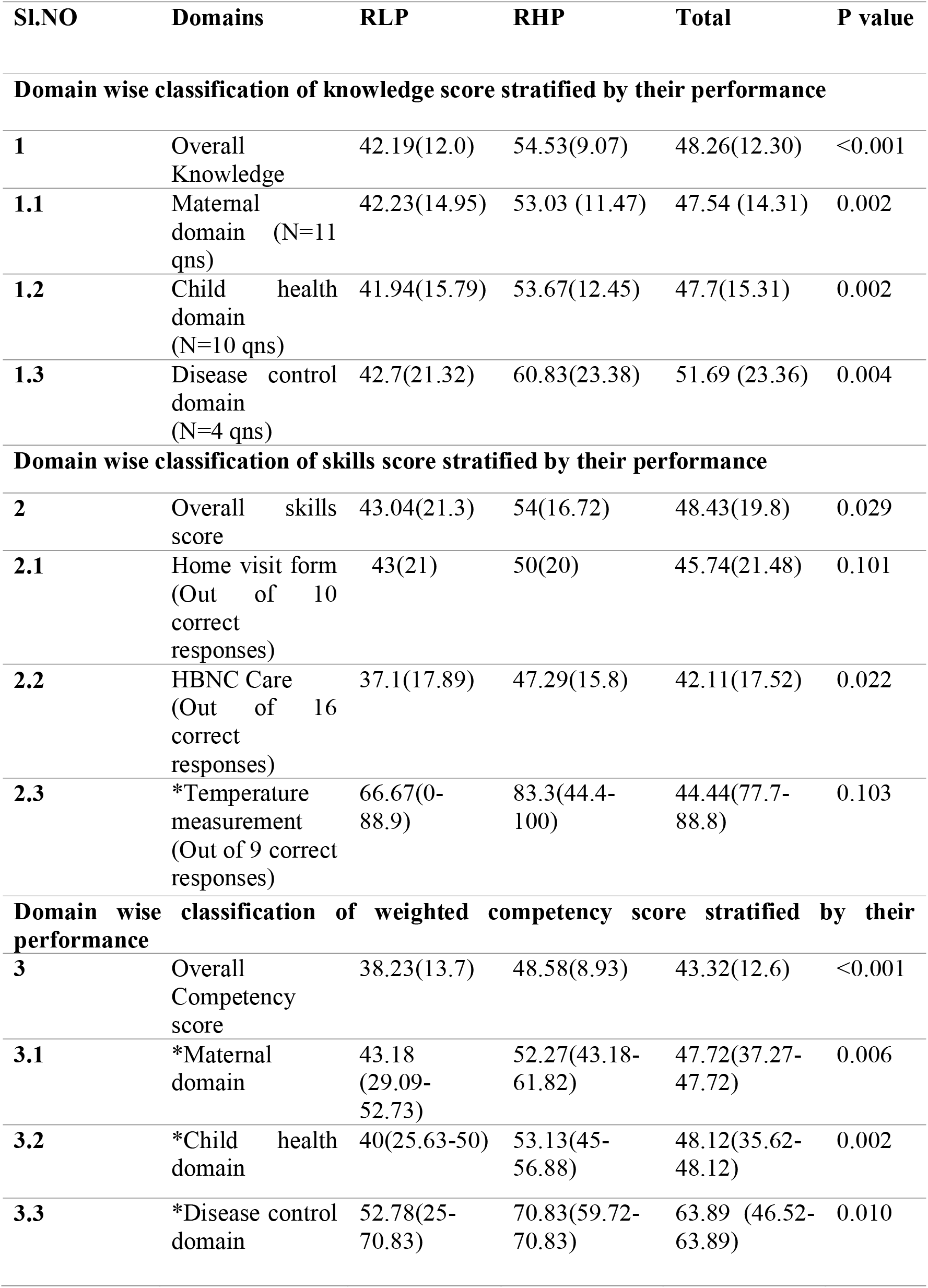
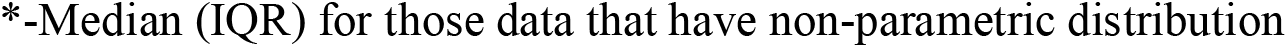
Domain wise classification of knowledge, skills and the competency score stratified by their performance.

| Sl.NO | Domains | RLP | RHP | Total | P value |
| --- | --- | --- | --- | --- | --- |
| **Domain wise classification of knowledge score stratified by their performance** |  |  |  |  |  |
| **1** | Overall Knowledge | 42.19(12.0) | 54.53(9.07) | 48.26(12.30) | <0.001 |
| **1.1** | Maternal domain (N=11 qns) | 42.23(14.95) | 53.03 (11.47) | 47.54 (14.31) | 0.002 |
| **1.2** | Child health domain (N=10 qns) | 41.94(15.79) | 53.67(12.45) | 47.7(15.31) | 0.002 |
| **1.3** | Disease control domain (N=4 qns) | 42.7(21.32) | 60.83(23.38) | 51.69 (23.36) | 0.004 |
| **Domain wise classification of skills score stratified by their performance** |  |  |  |  |  |
| **2** | Overall skills score | 43.04(21.3) | 54(16.72) | 48.43(19.8) | 0.029 |
| **2.1** | Home visit form (Out of 10 correct responses) | 43(21) | 50(20) | 45.74(21.48) | 0.101 |
| **2.2** | HBNC Care (Out of 16 correct responses) | 37.1(17.89) | 47.29(15.8) | 42.11(17.52) | 0.022 |
| **2.3** | *Temperature measurement (Out of 9 correct responses) | 66.67(0-88.9) | 83.3(44.4-100) | 44.44(77.7-88.8) | 0.103 |
| **Domain wise classification of weighted competency score stratified by their performance** |  |  |  |  |  |
| **3** | Overall Competency score | 38.23(13.7) | 48.58(8.93) | 43.32(12.6) | <0.001 |
| **3.1** | *Maternal domain | 43.18 (29.09-52.73) | 52.27(43.18-61.82) | 47.72(37.27-47.72) | 0.006 |
| **3.2** | *Child health domain | 40(25.63-50) | 53.13(45-56.88) | 48.12(35.62-48.12) | 0.002 |
| **3.3** | *Disease control domain | 52.78(25-70.83) | 70.83(59.72-70.83) | 63.89 (46.52-63.89) | 0.010 |

\*-Median (IQR) for those data that have non-parametric distribution

Three models of multivariate logistic regression with socio-demographic factors as independent predictors and RLP/RHP as dependent variables were computed: in the first model, only individual factors were considered. Subsequently, in the second model, all the individual factors and immediate systemic factors were considered except competency score and in the final model all three were considered for multivariate analysis. All the three models revealed that for every 1 unit increase in age, there was 32% higher chance of better performance (OR: 1.32, CI:1.14-1.59, *p<0*.*001*) and those with educational qualification more than primary education were found to have 6.52 times (OR:6.52, CI:1.52-27.83, *p=0*.*011*) of better performance compared to those with primary education following adjustment of the confounders.

A 4*4 matrix containing 4 quadrants was constructed to establish relationship between age, education, competencies and the incentivised real-world performance as shown in Figure 1. Majority of the RHP CHWs over 30 years of age and above primary education had better theoretical competencies as compared to RLP CHWs. There were instances where few RHP CHWs over 30 years and upto primary level of education had higher competencies which might have been attributed by their work experience. While above primary education had contributed to better competencies in some RHP CHWs less than 30 years.

**Figure 1:**
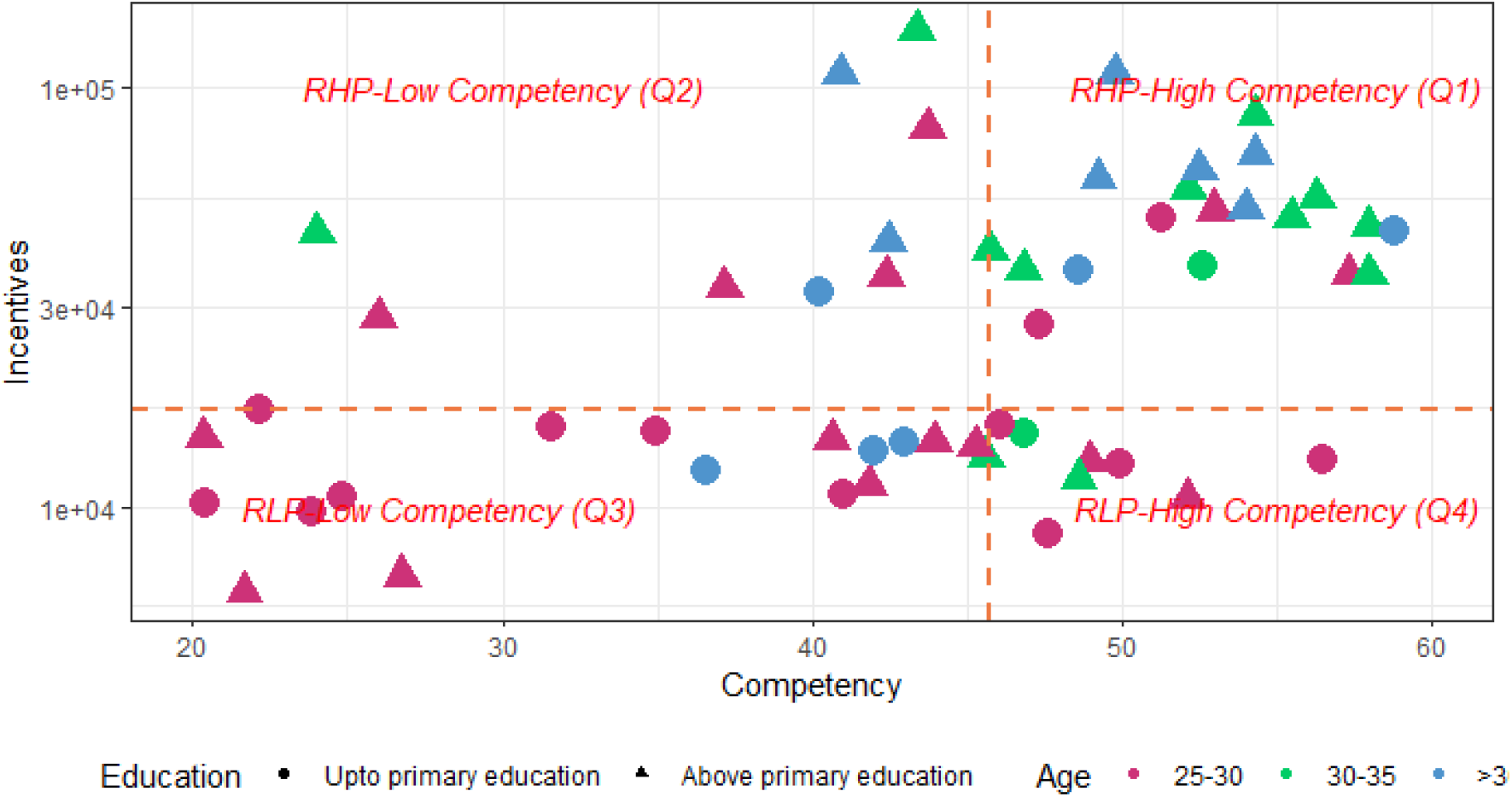
4*4 matrix on relationship between competencies and real-world performance.

Further cluster analysis, a multivariate method was employed to identify the number and nature of clusters formed among 61 CHWs based on characteristics like age, caste, education, population, distance and competency score the results of which are explained in supplementary file (Supplementary File 3). There were 2 clusters formed using two step cluster method with the overall model being fair and there was significant difference between the clusters and the performance (*P value =0*.*008*).

## Discussion

Majority of the CHWs were over 25 years of age, married and more than half of them belonged to below poverty line, Other backward classes with educational qualification upto primary education and more than five years of experience in the field. These findings were concordant with study ^[7]^ where only higher education was found to be associated with a better performance unlike factors like years of experience, age, marital status and social class which were found to have a mixed effect on performance. ^[13-17]^ Another study ^[18]^ shows increased years of experience was found to be associated with better performance and documented predominant involvement of married women belonging to below poverty line similar to our findings yet not significant. As their years of experience as CHW proportionally increases with age, their performance tends to improve due to community trust, supportive supervision and self-efficacy.

Overall, our study focussed on measuring the indirect output measures of CHW performance like knowledge, skills and competencies. Majority of the RHP CHWs had above average knowledge and skills on assessment as compared to RLP CHWs, the difference of which were statistically significant. However, overall knowledge and skills of the selected CHWs were below average. Colvin CJ et.al .2013 ^[19]^ quotes that lack of skills of the CHW may lead to lack of compliance with their health advice to community and ultimately loss of community trust. The possible explanations for better performance in RHP CHWs might be attributed to more than primary education and increased work experience relatively more than RLP CHWs under study.

The availability of drugs were better among the RLP CHWs than the RHP CHWs whereas equipments availability were vice-versa and all of them had the required registers. The probable reason could be the population, where RLP CHWs covered lesser population as compared to RHP CHWs . Also, to some extent it depends on the procurement and distribution of the drugs by their supervisors and the logistics flow mechanism which needs to be explored qualitatively. This finding is contradictory to studies ^[20,21]^ which have reported shortage of drug and equipment supplies.

There is a direct relationship between competencies and performance influenced by the effect modifiers like age and education. Majority of the RHP CHWs over 30 years of age and above primary education had better theoretical competencies as compared to RLP CHWs. Further qualitative enquiry is necessary to understand the factors that might have influenced the RLP CHWS less than 30 years and upto primary education to achieve better competencies despite their educational and work experience gap.

## Conclusion and recommendations

CHWs being an intermediary link between the health system and the community, the relationship between their truest competencies and performance based incentivisation tends to have been influenced by age and education. This differentiation could help the policy makers in developing a more focused supportive supervision catering to their needs.

## Data Availability

All data produced in the present study are available upon reasonable request to the authors

## Abbreviations

ASHAs: Accredited Social Health Activists
CHC: Community Health Centre
CHWs: Community Health workers
IQR: Interquartile range
NHM: National Health Mission
ODK: Open Data Kit
ONA: Organisational network analysis
PHC: Primary Health Centre
RHP: Relatively High Performing
RLP: Relatively Low Performing
XLS: excel Spreadsheet.

## Availability of data and material

Data will be made available to researchers on reasonable request to the corresponding author.

## Acknowledgements

Permission and facilitation for data collection at field sites were provided by Block Medical Officer, Obedullaganj block, Raisen District, Madhya Pradesh.

Our sincere thanks to Dr. Deepti Dabar for her valuable guidance and feedback during the process of protocol writing and submission.

We thank our Head of the department, Prof (Dr.) Arun Kokane for his support that had helped us to mobilize the CHWs during our study at various settings.

We thank all the Senior residents, Junior Residents and the interns who helped us during the data collection period and for their cooperation during the study.

---

**Supplementary Table 1:**
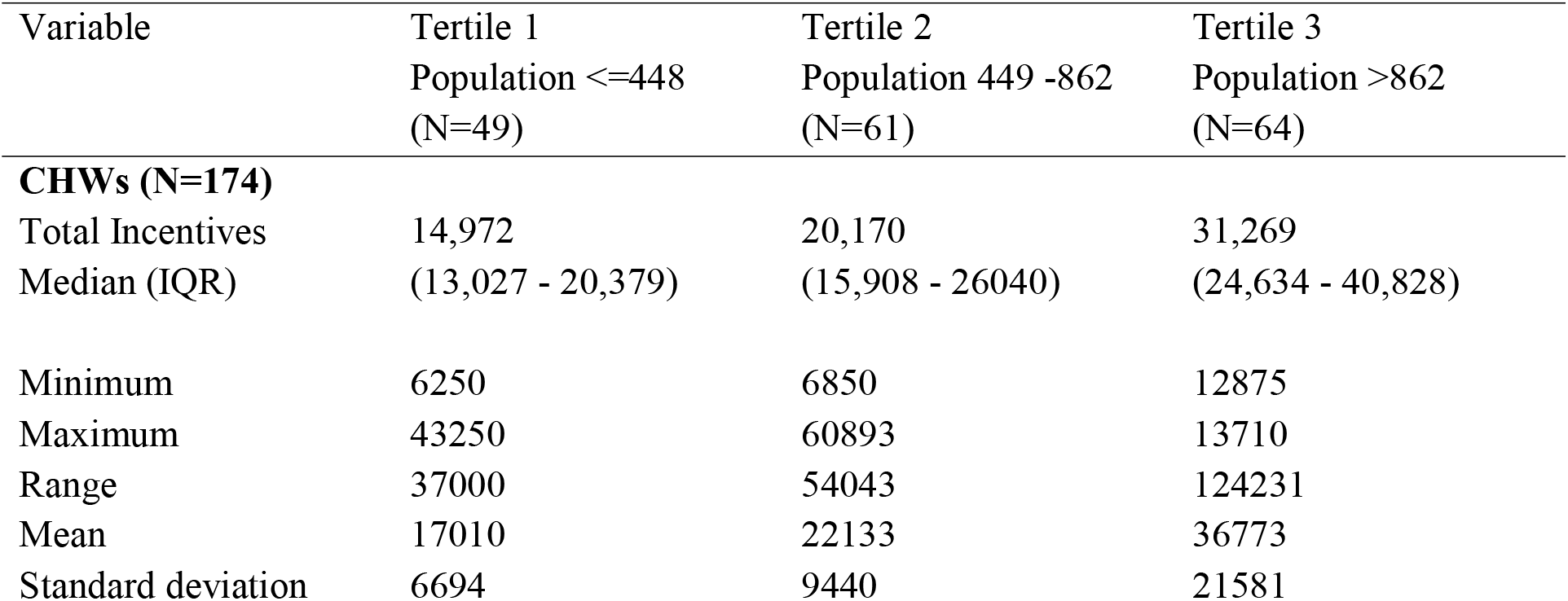
Distribution of overall incentives stratified by the population tertiles (N= 174)

**Supplementary Figure 1:**
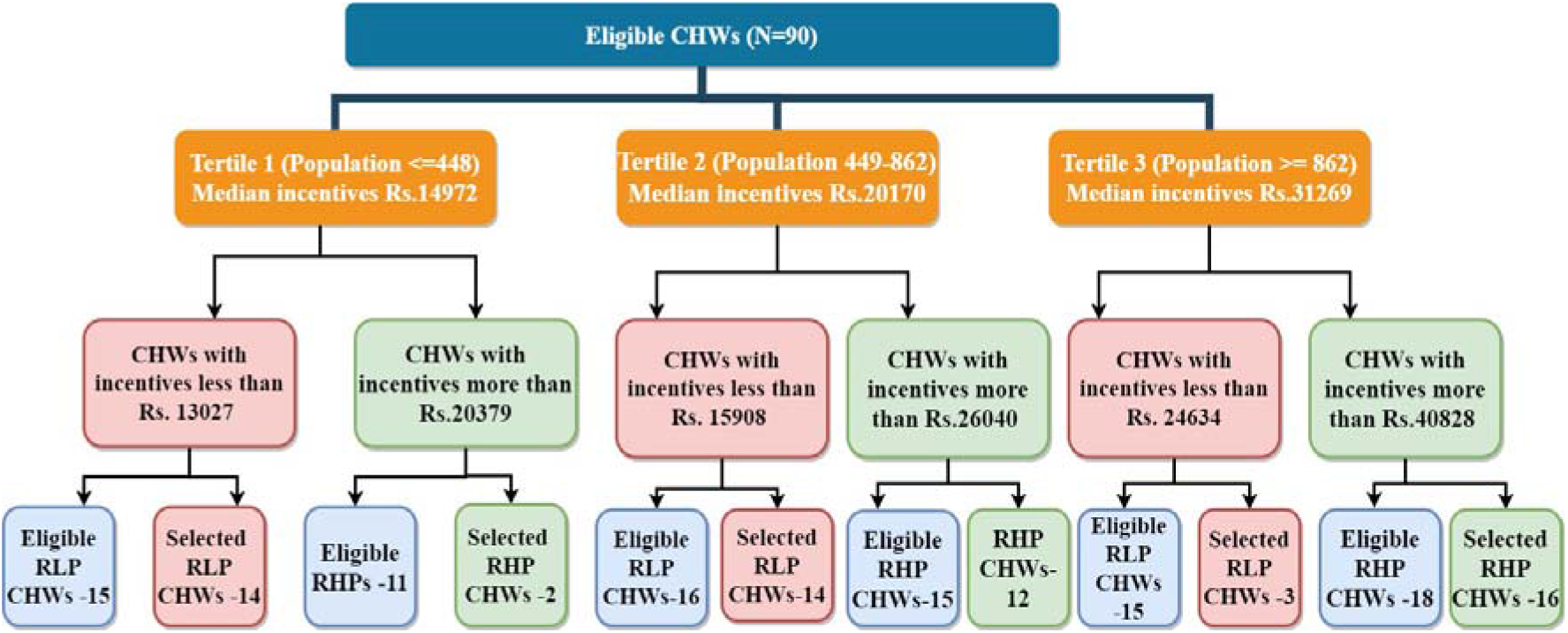
Flowchart showing final number of interviewed CHWs :

## Additional information

### A. Supplementary Files

1. Supplementary File 1: PBI (Performance based incentives system) CHWs
2. Supplementary File 2: Study Questionnaire.
3. Supplementary File 3: Cluster analysis

