## Supplementary File 1 for "An exploration of attributes effecting the relationship between competencies and performance of CHWs in a rural block"

A total of 22 variables has been enlisted with honorarium (incentive) pertaining to performance in various domains like maternal and child health e.g., Rs.150 for motivating PPIUCD insertion per case etc.

To note: Incentives are provided according to Indian currency (Rupees)

**Supplementary Table 1: Performance Based incentives of CHWs of Madhya Pradesh India**

| Incentives No | Activities performed | Incentives proposed |
| --- | --- | --- |
| 1.1.1 | Village survey | 300 |
| 1.1.2 | Birth registration | 300 |
| 1.1.3 | Marriage registration | 300 |
| 1.4 | Pregnancy registration | 300 |
| 1.5 | Listing beneficiaries for routine immunisation (Under 5) | 300 |
| 1.6 | VHND | 150 |
| 1.7 | PHC/CHC meeting | 350 |
| 2 | Pregnancy reg, Folic acid & albendazole | 200 |
| 3 | At least 4 ANC visits | 600 |
| 4 | Iron sucrose infusion for ANW with anaemia | 400 |
| 5 | MTP | 150 |
| 6 | HBNC | 250 |
| 7 | SNCU - Follow up of discharged children 4 times | 200 |
| 8 | Referral to NRC | 800 |
| 9 | Fully immunised (1 year) | 100 |
| 10 | Complete immunisation (2 year) | 75 |
| 11 | Complete immunisation (5-6 year) | 50 |
| 12.1 | Tubectomy motivation | 300 |
| 12.2 | Vasectomy motivation | 400 |
| 12.3 | PPIUCD insertion | 150 |
| 12.4 | PPIUCD insertion following abortion | 150 |
| 12.5 | Tubectomy within 7 days post delivery | 300 |
| 13.1 | Motivation for 1st conception by two years following marriage | 500 |
| 13.2 | Motivation for interpregnancy interval by three years | 500 |
| 13.3 | Tubectomy by two child norm | 1000 |
| 14 | Strengthening Mother and daughter in law relationships through campaigns | 100 |
| 15 | ASHA Kit for newlywed couples | 100 |
| 16 | Eligible couple survey | 250 |
| 17 | Antara dose injection | 100 |
| 18.1 | Malaria slides preparation | 15 |
| 18.2 | Radical treatment for Falciparum malaria | 75 |
| 18.3 | Testing for Plasmodium falciparum malaria | 75 |
| 19.1 | Identification of leprosy cases | 250 |
| 19.2 | Paucibacillary treatment | 400 |
| 19.3 | Multibacillary treatment | 600 |
| 20.1 | DOTS provider for newly diagnosed Tb cases | 1000 |
| 20.2 | DOTS provider for the relapse case | 1500 |
| 20.3 | DOTS provider for MDR Tb case | 5000 |
| 21 | Verbal autopsy of any 15–49-year-old women within 48 hrs of death | 200 |
| 22 | Gestational diabetes testing in pregnant women | 800 |
