## Supplementary File 2 for "An exploration of attributes effecting the relationship between competencies and performance of CHWs in a rural block"

A. **Knowledge assessment of CHWs:**

**Instructions to be followed:**

- *Read the following questions properly and answer all the questions by choosing the*
- *The appropriate answer with a tick mark.*
- *Kindly tick only one among the following options.*
- *Do not overwrite.*
- *You will be provided with 15 minutes time to answer the following questions.*

1.Which is the ideal period of early registration?

a. 10-12 weeks	c. 12-16 weeks
b. 12-14 weeks	d.14-20 week

2.What are the potential danger signs during pregnancy?

a. Any vaginal bleeding during pregnancy
b. Severe headache / Blurring of vision
c. No perception of fetal movements
d. All the above

3. All the following are the 5C’s of pregnancy except?

a. Clean	c. clean surface
b. Clean cord stump	d. clean thread

4.Which of the following is not the danger sign of post-natal period?

a. Fever	c. excessive vaginal bleeding

b. severe abdominal pain	d. vomiting

5. what are not the signs of referral to the first referral unit?

a. poor breast sucking	c. fast breathing

b. blue palms and soles	d. no meconium passed in 48 hrs.

6. Termination of pregnancy is legal till?

a. 14 weeks	c. 16 weeks

b. 18 weeks	d. 20 weeks

7. The following are the golden rules to be followed in a child with diarrhea except?

a. Continue feeding	c. warmth

b. Give ORS	d. Refer in case of danger signs

8. Emergency contraceptive pill must be taken within?

a. 24hrs	c. 36hours

b .48 hours	d. 72 hours

9. HIV is transmitted by all of the following except?

a. sexual route	c. blood

b. vertical transmission	d. sharing other utility items

10. All the following are the premenstrual syndrome except?

a. sore breast	c. Feeling extra tired

b. constipation	d. vomiting

11. All the following are the presumptive symptoms of tuberculosis except?

a. persistent cough> 2 weeks duration	c. high grade fever

b. weight loss	d. occasional hemoptysis

12. Number of chloroquine 150mg base Tablets to be given in an adult > 15 years old?

a. 6	c. 5

b. 4	d. 3

13. When do you consider the child completely immunized for age? if child has received

a. BCG+ Pentavalent 1,2,3 doses + OPV 1,2,3 + Measles at 12 months	b. BCG+ Pentavalent 1,2,3 doses + OPV 1,2,3 at 12 months	C. BCG+ Pentavalent 1,2,3 doses + OPV 1,2,3+ Measles at 10 months

d. BCG+ Pentavalent 1,2,3 doses + OPV 1,2,3 at 10 months

14. Given the last menstrual period of a women to be 1/1/2018. what is the expected delivery Date?

a. 8/10/2018	c. 7/ 10/2018
b. 9/9/2018	d. 8/9/2018

15. Who are the ideal candidates of Copper T insertion?

a. Those who had at least one child

b. Those who don’t have access to health care facility

c. Those with menstrual problems

d. None of the above

16. According to the HBNC guidelines, how many number of visits should be done in case of home deliveries?

a. 8	c. 7

b. 6	d. 5

17. Iron Tablets are initiated in pregnancy after?

a. Soon after confirmation of pregnancy	c. 2 months onwards

b. 3 months onwards	d. 5 months onwards

18. How many tetanus injections doses are given in a primi pregnancy?

a. 2	c.1

b. 3	d. none

19. Following are the danger signs of diarrhea in a child except?

a. blood in stools	c. lethargy

b. unable to drink or breast feed	d. fever

20. what is the temperature above which fever is diagnosed?

a. 42-degree	b. 39 degree

c. 40-degree	d. 36 degree

21. The following are the grounds of medical termination of pregnancy except?

a. When continuing pregnancy causes a risk to the life of a pregnant mother

b. when there is a substantial risk that if a child is born would be seriously handicapped

due to physical or mental abnormalities

c. When pregnancy is caused due to rape.

d. when there is an issue of unwanted pregnancy.

22. What is the hemoglobin above which the pregnant woman is said to have no anemia?

a. 12 g/dl	c.11g/dl

b. 13g/dl	d.10g/dl

23. How much breadths above the following is considered as fast breathing in a child with 1year age?

a.50 breadths/min	c. 30breadths/min

b. 40 breadths/min	d. 25breadths/min

24. what is the dose of albendazole that has to be given in a 2year child?

a. ½ Tablet of 400mg	c. 1 Tablet of 400mg

b. ½ Tablet of 200mg	d. 1 Tablet of 200mg

25. How long should iron and folic acid Tablets been given in a child with anemia?

a. 30 days	c. 6 months

b. 5 months	d. 14 days

**B. Skill assessment of the CHWs:**

***Instructions to be followed:***

- Make sure that the prerequisites for the skill observation are readily available.
- Use the checklist while observing the skills being implemented.
- When a step is performed correctly place a tick mark in the box corresponding to it.
- When a step is not performed correctly, place a cross mark in the box.
- Inform CHW to perform that task in their step wise manner
- For the correct response 1 mark will be awarded and for the incorrect response no marks.
- No specific time limits

**Supplementary Table 3: Home Visit Form (Examination of Mother and Newborn)**

| Ask /Examine  Date of CHW's visit | Supervisory check (Marks obtained) |
| --- | --- |
| Ask Mother |  |
| No. of times mother takes full meal in 24 hrs. |  |
| Bleeding: How many pads are changed in a day |  |
| During the cold season, is the baby being kept warm (near mother, clothed and wrapped properly) |  |
| Is baby crying incessantly or passing urine less than 6 times a day |  |
| Examination of mother |  |
| Temperature: Measure and record |  |
| Foul smelling discharge and fever more than 100-degree F (37.8 degree C) |  |
| Is mother speaking abnormally or having fits? |  |
| Mother has no milk since delivery or if perceives breast milk to be less |  |
| Cracked nipples/painful and/or engorged breast |  |
| Examination of the baby |  |
| Are the eyes swollen or with pus |  |
| Weight (on day 7, 14, 21 and 28) |  |
| Temperature: Measure and Record |  |
| Skin:  Pus filled pustules |  |
| Cracks or redness on the skin fold (thigh/Axilla/Buttock) |  |
| Yellowness in eyes or skin: Jaundice |  |

***Supplementary Table 4: Home based neonatal care - checklist:***

|  | Checklist | | Mark obtained |
| --- | --- | --- | --- |
| 1 | Did the CHW wash her hands before examining the child? | |  |
| 2 | Did the CHW measure the weight of the child | - ‘0’ adjusted before start of weight |  |
|  |  | - measured the weight |  |
| 3 | Did the CHW plot weight of the child on the MCP card | - Plotted the growth chart – markings made in green/yellow/red colour zone |  |
|  |  | - ASHA recorded child’s weight in her register |  |
| 4 | Did the CHW check the immunization status of the child | - Checked the MCP card |  |
|  |  | - Informed mother about the next dose of the vaccine |  |
| 5 | a. Did the CHW measure the temperature of the child using thermometer? | |  |
|  | b. If thermometer is not available, did the CHW place her hand on child’s tummy to decide if the child feels hot to touch | |  |
| 6 | Did the CHW check for breathing counts | - Made sure the child was quiet and calm. |  |
|  |  | - Removed the clothes of the child gently to see the moving chest. |  |
|  |  | - According to watch counted the child’s breathing for one minute. |  |
| 7 | Did the CHW check for umbilical cord care | - checked the cord and its surrounding skin without touching it |  |
| 8 | Did the CHW assess the hydration status of the child | - pinched the skin of the child over the abdomen |  |
| 9 | Did the CHW check the skin of the child | - checked for any pus –filled pustules anywhere on the skin of the child |  |
| 10 | Did the CHW check the eyes of the child | |  |
| 11 | Did the CHW check for yellowness/Jaundice | - Pressed the forehead or nose of the child gently and checked for the yellowness |  |
| 12 | Did the CHW look for stiffness in neck | - Look to see if the child moves and bents his neck easily as he look around |  |

**Supplementary Table 5: Temperature assessment checklist**

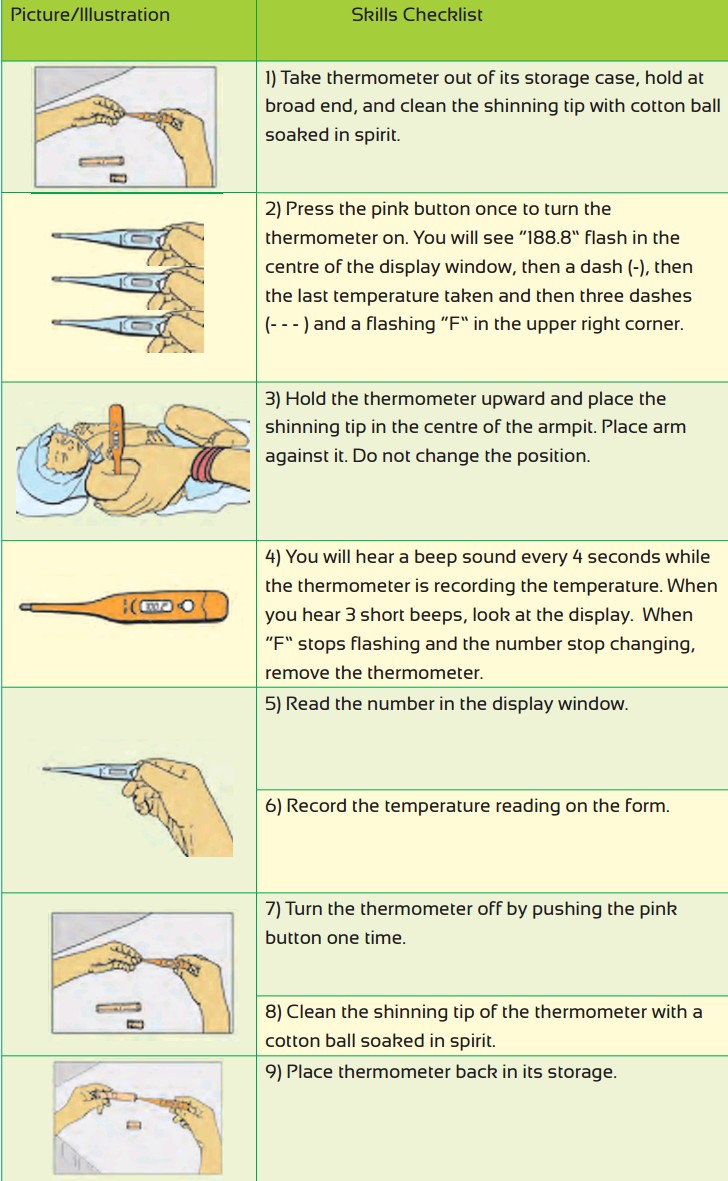

**Supplementary Table 6: Checklist for the drugs, equipments and registration forms among CHWs:**

**Instructions:**

The following drugs, equipments and the registers that are listed must be verified for the availability with the concerned CHW.

If the drug, equipment and the register form is available, kindly tick in the corresponding box and if not available put cross in the concerned box.

| ***Drugs availability*** | ***Status of availability*** |
| --- | --- |
| Tab paracetamol |  |
| Paracetamol syrup |  |
| Tab Iron Folic Acid |  |
| Tab Dicyclomine |  |
| Tetracycline ointment |  |
| Zinc Tablets |  |
| Povidine Ointment Tube |  |
| G.V.Paint |  |
| Cotrimoxazole syrup |  |
| Paediatric Cotrimoxazole Tablets |  |
| ORS packets |  |
| Condoms |  |
| Oral pills (in cycles) |  |
| Spirit |  |
| Soap |  |
| Sterilized Cotton |  |
| Bandages, 4 cm* 4 meters |  |
| Nischay kit |  |
| Rapid diagnostic kit |  |
| Slides for Malaria & Lancets |  |
| Emergency Contraceptive Pill |  |
| Sanitary napkins |  |
| **Equipments availability:** |  |
| Digital wrist watch |  |
| Thermometer |  |
| Weighing Scale for new born |  |
| Baby blanket |  |
| Baby feeding spoon |  |
| Kit bag |  |
| Communication Kit |  |
| Mucus Extractor |  |
| **Forms available:** |  |
| CHW drug kit stock card |  |
| Format for individual plans (birth preparedness) |  |
| Delivery form |  |
| First examination of the newborn |  |
| Home visit form |  |
| **Records available:** |  |
| *Village health register* |  |
| *ASHA diary* |  |
