## Supplementary File 3 for "An exploration of attributes effecting the relationship between competencies and performance of CHWs in a rural block"

**Supplementary Figure 2: displaying the cluster model of the selected variables (N=61)**

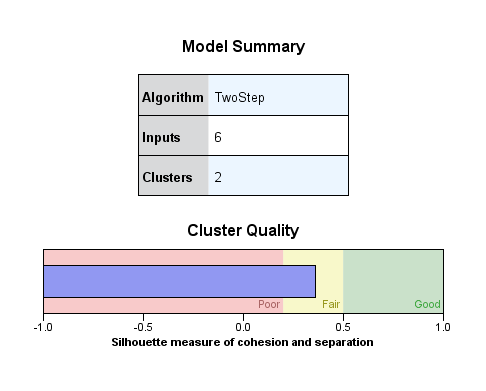

**Supplementary Table 7** displays the characteristics of each of the cluster based on our 6 inputs where most of the CHWs i.e., 35 belonged to cluster 1, the details of the characteristics has been described below for each cluster.

**Supplementary Table 8: shows the descriptive analysis of the clusters ( N=61)**

| Characteristics | Cluster 1 | Cluster 2 |
| --- | --- | --- |
| Number of CHWs in each cluster | 35 | 25 |
| Mean Age (years) | 31.43 | 30.4 |
| Education | All had been educated above primary level | All had been educated up to primary level |
| Caste | 48.6% of ASHAs belonged to OBC | 36% ASHAs belonged to Scheduled tribe |
| Mean Competency score(%) | 46.03% | 39.27% |
| Population | 1137.54 | 532.36 |
| Distance | 9.70 | 11.78 |

**Supplementary Figure 3** below shows the cluster size in a pie chart below and the ratio of smallest (cluster 2) to the largest cluster ( cluster 1) of 1.4

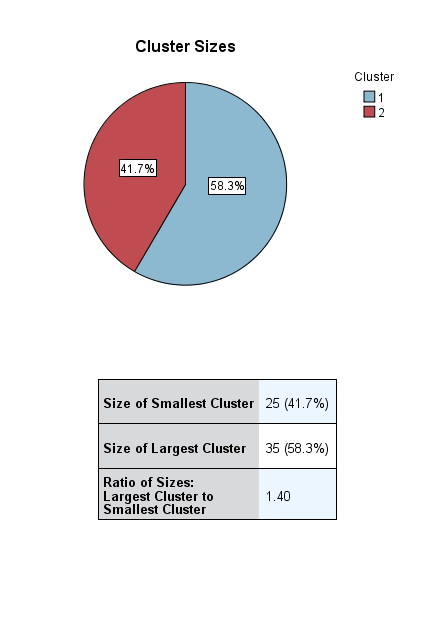

A cross tabulation was made as shown in **Supplementary Table 9** to find the association between the two clusters and the performance using chi square and the pattern of stratification of performance between them. It was found that there was significant difference between the clusters and the performance ( P value 0.008).

**Supplementary Table 9**: **Distribution of clusters stratified by the performance**

| **Performance** | **RLP** | **RHP** |  | **Unadjusted odds ratio** | **95% Confidence interval** | **P value** |
| --- | --- | --- | --- | --- | --- | --- |
| Cluster 1 | 13 | 22 |  | 4.3-52 | 1.43-13.21 | 0.009 |
| Cluster 2 (ref) | 18 | 7 |  | - | - | - |
| Total | 31 | 29 |  |  |  |  |

Further we had performed univariable logistic regression for the same, the results of which are displayed in **Supplementary Table 9.** It was found that those CHWs belonging to Cluster 1 had 4.32 times higher odds of better performance (P value 0.009) as compared to Cluster 2 .
